# Immediate postoperative minimal residual disease detection with MAESTRO predicts recurrence and survival in head and neck cancer patients treated with surgery

**DOI:** 10.1101/2025.01.27.25321202

**Authors:** Edward S. Sim, Justin Rhoades, Kan Xiong, Laurel Walsh, Andjela Crnjac, Timothy Blewett, Yana Al-Inaya, Julia Mendel, Daniel A. Ruiz-Torres, Vasileios Efthymiou, Gjystina Lumaj, William J. Benjamin, G. Mike Makrigiorgos, Shervin Tabrizi, Viktor A. Adalsteinsson, Daniel L. Faden

## Abstract

**Purpose:** While circulating tumor DNA (ctDNA) is a promising biomarker for minimal residual disease (MRD) detection in head and neck squamous cell carcinoma (HNSCC), more sensitive assays are needed for accurate MRD detection at clinically-relevant timepoints. Ultrasensitive MRD detection immediately after surgery could guide adjuvant therapy decisions, but early ctDNA dynamics are poorly understood.

**Experimental Design:** We applied MAESTRO, a whole-genome, tumor-informed, mutation-enrichment sequencing assay, in a pooled testing format called MAESTRO-Pool, to plasma samples from HNSCC patients collected immediately after surgery and during surveillance. We evaluated whether early MRD detection could predict outcomes.

**Results:** Among 24 predominantly HPV-independent (95.8%) HNSCC patients, rapid ctDNA clearance occurred by the first postoperative sample (1-3 days postoperatively) in 9 patients without an event (recurrence or death). 13/15 patients with an event were MRD+ (PPV = 92.9%; NPV = 80%) with a median tumor fraction (TFx) of 54 ppm (range 6-1,177 ppm). In the first and last sample of the immediate postoperative window, 8/13 and 10/13 patients had TFx below 100 ppm, respectively, the detection limit of leading commercial assays. Early MRD detection correlated with worse overall survival (HR = 8.3; 95% CI: 1.1-66.1; *P* = 0.02) and event-free survival (HR = 27.4; 95% CI: 3.5-214.5; *P* < 0.0001) independent of high-risk pathology.

**Conclusions:** Immediate postoperative MRD detection by MAESTRO was predictive of recurrence and death. Given the ultralow TFxs observed, ultrasensitive assays will be essential for reliable MRD detection during early postoperative timepoints to enable personalized adjuvant therapy decision-making in HNSCC.

## Introduction

Most patients with head and neck squamous cell carcinoma (HNSCC) present with locoregionally advanced disease (1). Despite standard-of-care treatments with surgery followed by adjuvant radiation or chemoradiation, survival outcomes for HNSCC patients remain poor, particularly for HPV-independent (tobacco/alcohol-related) disease with recurrence rates of at least 50% (2). When undergoing curative-intent surgical resection, patients can receive no additional treatment, adjuvant radiation therapy alone, or combination chemoradiotherapy for high risk patients. Ongoing studies are evaluating the addition of immunotherapy as well for the highest risk patients to further escalate treatment in hopes of improving survival (3). The decision-making process for selecting adjuvant treatment options is guided by clinicopathologic risk factors. According to the current guidelines from the National Comprehensive Cancer Network, patients with either extracapsular extension or positive margins on surgical pathology are advised to undergo combination chemoradiotherapy (1). However, such stratification is non-personalized and imperfect, as are the methodologies for assessing risk-factor status, such as the determination of surgical margins (4).

Following treatment, surveillance for recurrence relies on frequent physical exam and cross-sectional imaging. However, single-center prospective and retrospective studies have shown that imaging-based surveillance approaches do not improve locoregional control (LRC), disease free survival (DFS), or OS relative to surveillance without imaging (1,5–8). In addition, imaging-based paradigms often do not change clinical decision-making; one study estimated that 1108 scans would be required to detect one asymptomatic recurrence for which the scan would lead to successful salvage treatment (5,9). Imaging surveillance typically begins with re-staging scans 3 months after the end of definitive treatment, including adjuvant therapy. Three months is the earliest recommended imaging time due to the lack of specificity of current approaches (high rate of false positives) and thus poor positive predictive value of assessing disease status earlier after treatment (10). This is largely due to post-treatment changes in the resected tumor bed and draining lymph nodes, highlighting the need for better diagnostic tests to detect persistent or recurrent disease. A systematic review and meta-analysis of 3-month post-treatment PET/CT in HNSCC reported a positive predictive value (PPV) of only 58% (11).

Liquid biopsy tests that assay circulating tumor DNA (ctDNA) have arisen as a promising method of detecting minimal residual disease (MRD) when the tumor is below the detection level of clinical or radiologic tools (12–14). MRD detection through ctDNA can also be performed earlier after treatment, as the methodology is agnostic to the problems which disallow early imaging-based MRD detection. Prior studies of ctDNA in HNSCC have shown promise in diagnosing disease recurrence but have largely been restricted to HPV-associated oropharyngeal HNSCC with less robust data demonstrating the efficacy of ctDNA in HPV-independent disease (15–19). This is partly due to the difficulty of capturing the vast genomic heterogeneity seen in HPV-independent HNSCC (20). Additionally, early ctDNA kinetics in the immediate post-operative setting in HPV-independent HNSCC and its potential to serve as a reliable biomarker of persistent disease have not yet been studied. Identifying a reliable predictor of persistent disease before initiating adjuvant therapy could guide decisions on escalating or de-escalating adjuvant therapies, potentially improving patient outcomes and reducing morbidity and mortality.

While MRD detection by measuring ctDNA levels has shown promise, higher sensitivity is needed (21). For example, in a recent study of 100 HNSCC patients utilizing a leading tumor-informed commercial ctDNA assay, only 75% of patients had detectable ctDNA at baseline, prior to the start of treatment, a clinical scenario in which there is significantly more tumor burden than after treatment (MRD), highlighting the challenges of reliable MRD detection in HNSCC (22). To address this need, our group has developed MAESTRO (Minor Allele Enriched Sequencing Through Recognition Oligonucleotides), a method for assaying thousands of patient-specific tumor mutations, identified by whole-genome sequencing of a patient’s tumor and buffy coat, in cell-free DNA (cfDNA), with minimal sequencing. By using short allele-specific oligonucleotides under optimized thermodynamic conditions, MAESTRO enriches rare mutations and enables highly accurate and efficient ctDNA detection with up to 100-fold less sequencing (23,24). Given the minimal sequencing required, MAESTRO tests can also be pooled and applied to plasma samples from many patients (MAESTRO-Pool) allowing simultaneous detection of MRD in patient-matched samples while also assessing specificity in patient-unmatched samples (25–27). In this prospective observational cohort study, we evaluated the utility of MAESTRO-Pool for MRD detection in the immediate post-operative period, while patients were still in the hospital following surgery, a clinically relevant time window for adjuvant decision making in HNSCC.

## Methods

### Patients and Sample Collection

This was a single-center, non-interventional, prospective cohort study. All HNSCC patients evaluated at Massachusetts Eye and Ear (Boston, MA) between April 2022 and December 2023 undergoing definitive surgery were considered for this study. Patients with known distant metastasis or other active malignancies at the time of enrollment were excluded. Patients were prospectively enrolled in this study under Dana-Farber/Harvard Cancer Center Institutional Review Board (#18-653). All patients underwent preoperative staging imaging to exclude distant metastasis with computed tomography (CT) and/or a positron-emission tomography (PET). Written informed consent was obtained from each enrolled patient. This research was conducted in accordance with the provisions of the Declaration of Helsinki, and the U.S. Common Rule. Blood samples were collected using EDTA or cell-free DNA collection tubes (Streck, La Vista, Nebraska) preoperatively on the morning of surgery and serial samples were collected while patients were in the hospital recovering from surgery (post-operative window). Additional samples were collected at timepoints (surveillance period) during follow-up visits with providers. cfDNA was extracted and sequencing libraries were constructed as previously described (25–27).

### Clinical data collection and annotation

Trained clinical investigators reviewed electronic medical records for clinical data annotation. The following data were collected: age at diagnosis, biologic sex, smoking status, date of diagnosis and surgery, primary tumor site, tumor staging (American Joint Committee on Cancer Staging Manual [AJCC] eighth edition), surgical pathology features, and adjuvant and recurrence treatment modalities with corresponding start and end dates if applicable. Recurrence data, denoted as locoregional recurrence, distant metastasis or both, were abstracted from clinical visit notes as determined by confirmatory tissue biopsy. Vital status was recorded as either deceased, along with the corresponding date of death, or alive, with the date of the most recent follow-up visit noted. Event-free survival (EFS), defined as time from surgery to an event (disease progression, locoregional recurrence, distant metastasis or death), and overall survival (OS) were estimated using Kaplan Meier survival analysis. Patients without any event were censored at the date of last follow-up.

### MAESTRO tumor fingerprint design and data processing

MAESTRO-Pool assays were performed and data was processed as previously described (27). Briefly, whole genome sequencing of tumor and normal tissue was performed to identify patient-specific tumor somatic single nucleotide variants (SNVs) which were then used to design MAESTRO probes to track up to several thousand SNVs per patient. MAESTRO probes from all patients in the study were then pooled together into a MAESTRO-Pool panel which was then applied to all plasma samples. Tumor fraction, limit of detection (LoD95), and MRD status were determined as previously described (27). Tumor fraction represents the fraction of cfDNA fragments that are tumor derived. LoD95 reflects the minimal tumor fraction that would be estimated to be detected with 95% probability based on the number of cfDNA molecules and tumor mutations assayed in a given sample. Of note, LoD95 is not the lowest tumor fraction that MAESTRO can detect in a given sample. MRD was called using our dynamic MRD caller, which can calculate the probability of true tumor signal being detected in the data versus background noise based on sample-specific attributes such as observed tumor fractions, validated fingerprint sizes, and background SNV frequencies (27). For the primary analysis, we utilized the uniform background SNV frequency model for our dynamic MRD caller. We also used the sample-specific background SNV frequency model strictly to explore unmatched MRD detection in the cohort (26).

### Statistical Analysis

Log-rank testing was used to assess differences in event-free survival (EFS) and overall survival (OS) between patients with or without MRD. Unpaired 2-tailed t-tests with or without Welch’s correction were used to compare means of continuous variables. Cox proportional hazards regression modeling was used to control for potential confounding factors. All variables included in the multivariable Cox regression model met the proportional hazards assumption. All analyses were performed using R (v. 4.4.1; R Foundation).

## Results

### Patient Characteristics

We applied MAESTRO-Pool to a cohort of 24 locally advanced HNSCC patients who underwent curative-intent surgery. Patient and tumor characteristics are described in Table 1. The majority of patients had locally advanced disease (pathologic stage IV, n = 18, 75%), and none of the patients had evidence of distant metastases at time of surgery. The most frequent tumor subsite was the oral cavity (n = 19, 79%). One patient had p16-positive oropharyngeal SCC and the remainder were HPV-independent. As expected in a predominantly HPV-independent HNSCC population, patients were predominantly male (n = 18, 75%) and had a median age of 63 years (28). Eighty-three percent (n = 20) of patients had a primary tumor and the remaining had local recurrences or second primary cancers. Eighty-three percent (n = 20) of patients received adjuvant treatment with 33% (n = 8) receiving radiation only and 50% (n = 12) receiving chemoradiotherapy.

**Table 1:** Patient Characteristics

| Characteristic | Total Evaluated (N = 24) |
| --- | --- |
| **Age at Sugery, Median (IQR)** | 63 (60 – 70) |
| **Sex, n (%)** |  |
| Female | 6 (25) |
| Male | 18 (75) |
| **Smoking History, n (%)** |  |
| Current | 4 (17) |
| Former | 14 (58) |
| Never | 6 (25) |
| **Primary Subsite, n (%)** |  |
| Oral Cavity | 19 (79) |
| Larynx/Hypopharynx | 3 (13) |
| Oropharynx | 2 (8.3) |
| **Primary Tumor, n (%)** |  |
| Primary | 19 (79) |
| Second Primary or Recurrent | 5 (21) |
| **pT, n (%)** |  |
| pT1-pT2 | 4 (17) |
| pT3-pT4 | 20 (83) |
| **pN, n (%)** |  |
| pN0 | 8 (33) |
| pN1 | 1 (4.2) |
| pN2-pN3 | 15 (63) |
| **Overall Pathologic Stage, n (%)** |  |
| II | 2 (8.3) |
| III | 4 (17) |
| IVA-IVB | 18 (75) |
| **HPV, n (%)** |  |
| Negative | 13 (54) |
| Positive | 1 (4.2) |
| Not tested | 10 (42) |
| **Lymphovascular Invasion, n (%)** |  |
| Negative | 13 (54) |
| Positive | 11 (46) |
| **Perineural Invasion, n (%)** |  |
| Negative | 5 (21) |
| Positive | 19 (79) |
| **Extranodal Extension, n (%)** |  |
| Negative | 9 (38) |
| Positive | 7 (29) |
| NA (No Nodal Disease) | 8 (33) |
| **Margin Status, n (%)** |  |
| Negative | 21 (88) |
| Positive | 3 (13) |
| **Adjuvant Treatment, n (%)** |  |
| None | 4 (17) |
| Radiation only | 8 (33) |
| Chemoradiation | 12 (50) |

### ctDNA availability and MRD detection

All patients had detectable ctDNA at time of surgery with a median tumor fraction in cfDNA of 1,866 parts-per-million (ppm; range 159-134,843 ppm; Supp. Table 1; n=23, note: SK-47’s baseline plasma sample was lost due to an instrument error during cfDNA extraction). Serial blood draws were collected while patients remained in the hospital recovering from surgery (postoperative window) with a median of 3 samples per patient. The first postop draw occurred at a median of 1 day after surgery (range 1-3 days; Figure 1A) and all draws in the immediate postoperative window occurred within 10 days of surgery. Nearly all patients who were MRD negative during the postop period had undetectable ctDNA in the first postop blood draw following surgery (n = 10 patients) with the exception of SK-48 whose ctDNA became undetectable by their last postoperative draw, on Day 7 (Figure 1A).

**Figure 1:**
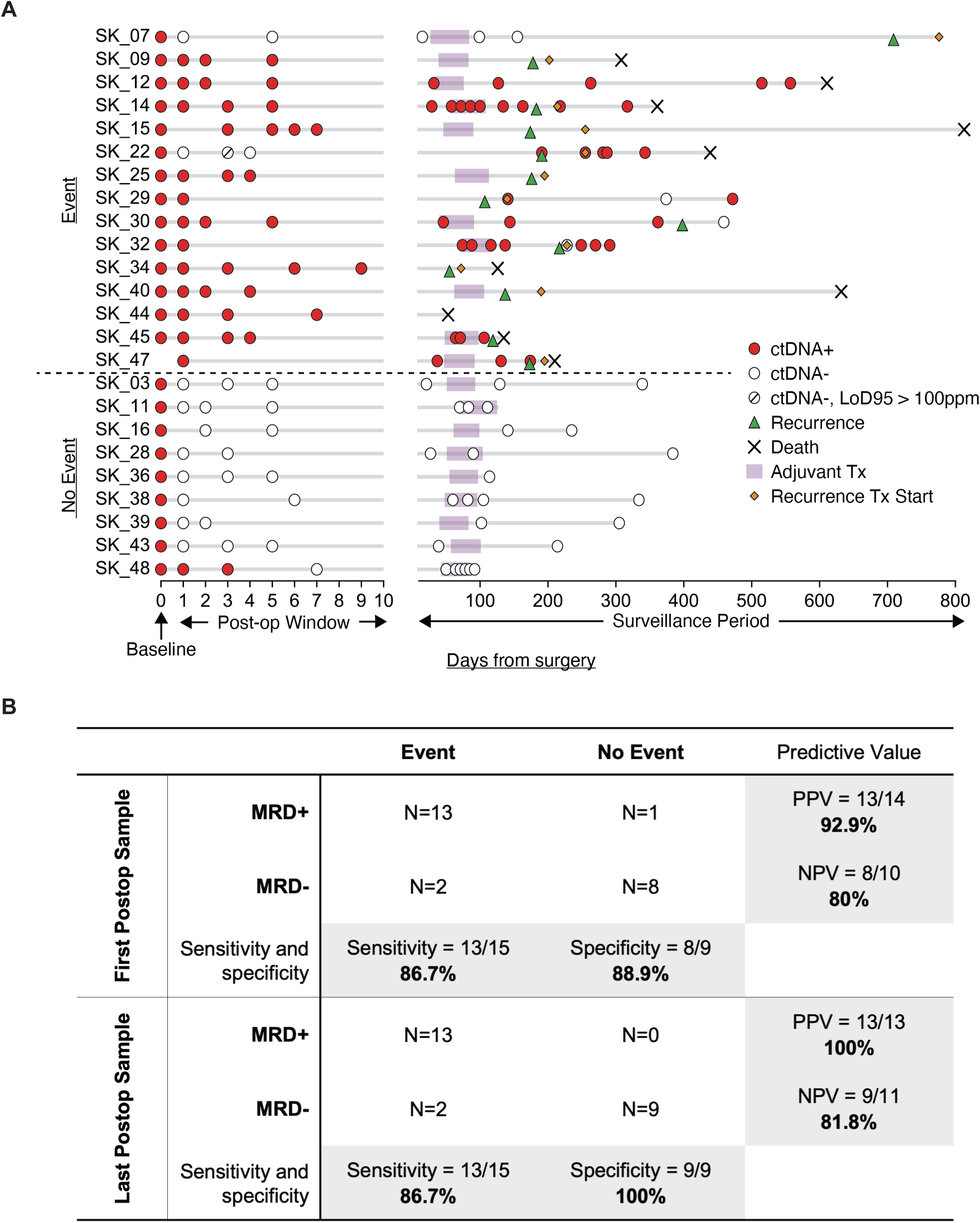
A) Swimmer’s plot of serial plasma sampling in the immediate postoperative setting and surveillance periods with corresponding MRD results. B) Diagnostic performance of MRD status in the first and last postoperative sample for predicting an event (recurrence or death).

MRD was detected in 14/24 patients (58.3%) on the first postoperative draw with a median tumor fraction of 51 ppm (range 6-1,177 ppm), and in 13/24 patients (54.2%) on the last sample during the postoperative window with a median tumor fraction of 23 ppm (range 2-214 ppm). The clinical sensitivity and specificity of MRD detection on the first postoperative blood draw for predicting an event (recurrence or death) was 86.7% and 88.9% respectively (Figure 1B). The clinical sensitivity and specificity of MRD detection on the last sample in the immediate postoperative window for predicting an event was 86.7% and 100% respectively (Figure 1B).

### MRD and survival

EFS and OS were compared for patients with detectable versus undetectable MRD on their first postoperative draw (within 1-3 days following surgery) and on the final draw within their postoperative window. The median follow-up for the entire cohort was 19.4 months (IQR: 9.5-22.5). MRD-negative patients had longer follow-up times (median = 20.3 months [IQR: 15.6-23.3] vs. median = 13.6 months [IQR: 7.8-22.1] for MRD-positive patients) due to deaths in the MRD-positive cohort. Patients who were MRD-negative on the first postoperative blood draw had significantly improved OS (HR = 7.4; 95% CI: 0.9-58.7; p = 0.03) and EFS (HR = 20.4; 95% CI: 2.6-159.6; p < 0.0001) compared to patients who were MRD-positive (Figure 2A). This strong survival benefit was also found when stratified by MRD status on the last postoperative blood draw (OS HR = 8.3; 95% CI: 1.1-66.1; p = 0.02 and EFS HR = 27.4; 95% CI: 3.5-214.5; p < 0.0001) (Figure 2B). A stepwise Cox multivariable regression model for event-free survival was developed using significant variables identified from univariable Cox regression analyses of *a priori* selected clinicopathologic features that may impact survival outcomes (Supp. Figure 1) (29). Due to a limited sample size, only the two most significant variables—MRD detection and extranodal extension and/or positive margins—were included in the final model. The final multivariable Cox regression model revealed that MRD was an independent predictor of EFS, outperforming established, clinically utilized pathological risk features used to guide adjuvant therapy escalation (Figure 2C, D)

**Figure 2:**
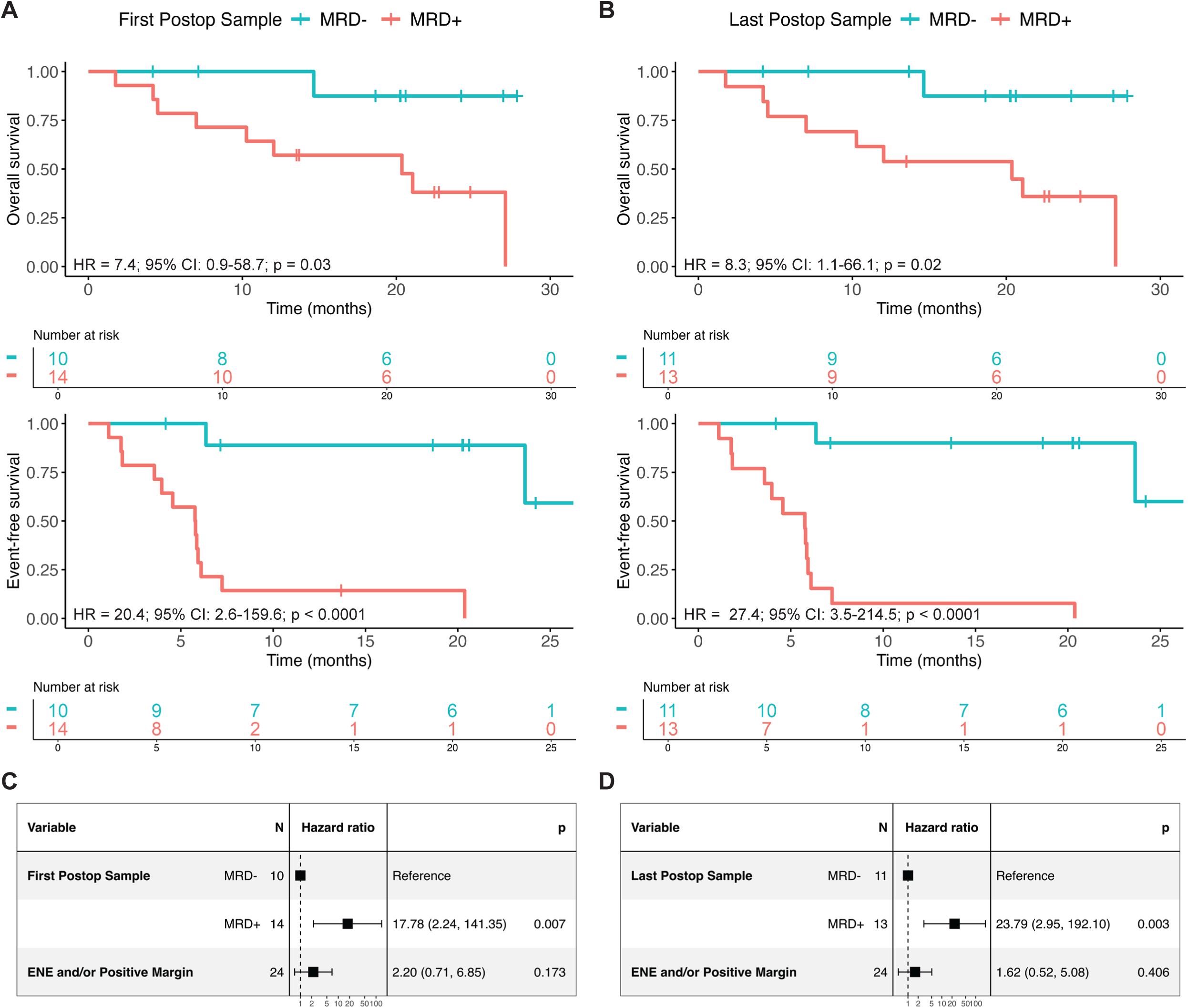
Overall survival, event-free survival Kaplan-Meier curves and multivariable Cox regression analyses for event-free survival of the 2 most significant variables from univariable Cox regression analysis when stratified by MRD status of the first (A & C) or last (B & D) postoperative sample. For Kaplan-curves, P values were obtained from log-rank testing with a significance cutoff of 0.05.

### Early ctDNA dynamics

We compared tumor fractions in cfDNA for the first and last postoperative blood draws in patients with and without events. In 8 of 13 patients (61.5%) with an event and detectable MRD in the first postoperative sample, tumor fractions were below the detection limit (100 ppm) of leading commercial assays (Figure 3A). This trend was even more pronounced in the last sample of the 10 day postoperative window, where 10 of 13 patients (77%) with an event and detectable MRD had tumor fractions below 100 ppm (Figure 3A). Of note, there were no statistical differences in limit of detection (p = 0.672) or number of mutations assayed (p = 0.135) between samples from MRD-negative and MRD-positive patients (Figure 3B). MAESTRO-Pool also allowed us to evaluate the specificity of each patient’s bespoke MAESTRO MRD test in unmatched samples from other patients. 3,523 of 3,565 (98.8%) of patient-unmatched tests were correctly called MRD negative (Supp. Figure 2A). This specificity increased to 3,553 (99.7%) with negligible impact on patient-matched MRD detection when sample-specific tuning (accounting for varied background SNV frequencies) was applied (Methods, Supp. Figure 2B-D) (26). When assessing early ctDNA dynamics within the postoperative window, tumor fraction levels decreased following surgery for all patients but patients with events tended to have tumor fractions that persisted at detectable levels during the postoperative period (Figure 3C, Supp. Figure 3). Most patients who did not have events had ctDNA clear by the first postoperative draw, as early as the first morning after surgery (Figure 3D, Supp. Figure 3).

**Figure 3:**
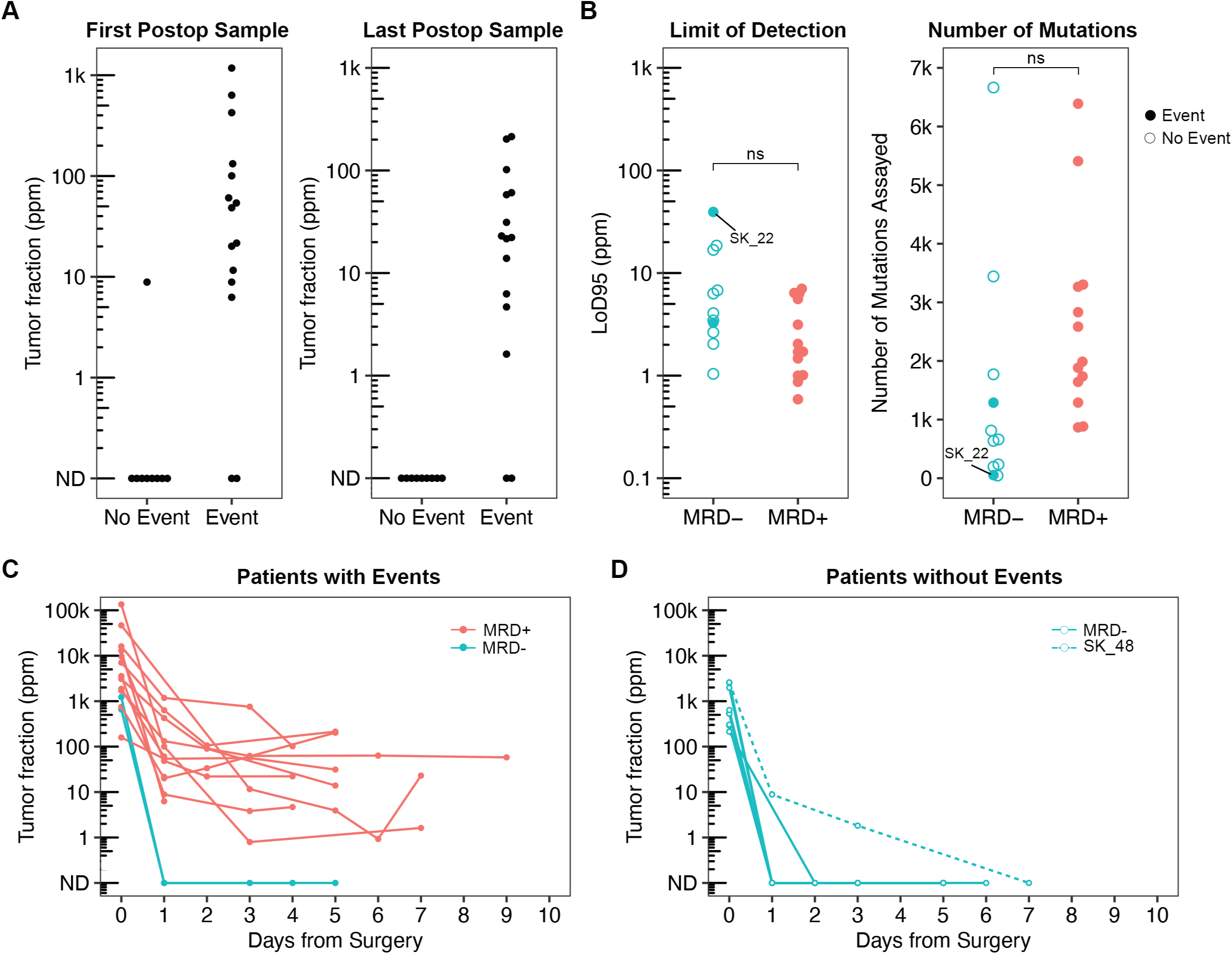
Tumor fraction dynamics in postoperative window. (A) tumor fraction levels in first and last postoperative samples in patients with vs without events. Each dot represents a single patient (B) Comparison of assay sensitivity in the last sample of the postoperative window for each MRD- vs MRD+ patient particularly assessing limit of detection (LoD95) in the left panel (p = 0.672) and fingerprint size (number of mutations assayed) in the right panel (p = 0.135). Number of mutations assayed in cfDNA were restricted to the number of mutations validated when MAESTRO was also applied to each patient’s tumor biopsy. Filled dots represent patients with events while unfilled dots represent patients without events. Red dots represent patients who were MRD+ while blue dots represent patients who were MRD-. SK_22 is labeled to demonstrate the assay sensitivity of a false-negative patient. (C) Early tumor fraction dynamics in patients who had events vs (D) no events. SK_48 is labeled to demonstrate the initially false-positive patient whose tumor fraction levels cleared gradually by day 7 after surgery. Statistical significance was assessed via Welch’s t-test in (B).

One patient (SK-48) was a false positive based on the first postoperative sample testing positive for MRD despite not having an event. However, by the last postoperative sample, the tumor fraction levels fell gradually to undetectable levels (Figure 3D, Supp. Figure 3). Two patients—SK_07 and SK_22—were false negatives in that no ctDNA was detected during the postoperative window but they eventually developed recurrence (Figure 1A). Patient SK-07 had pT3N0M0 squamous cell carcinoma of the left oral tongue and underwent partial glossectomy, left cervical lymphadenectomy, and reconstruction. The final surgical pathology revealed perineural invasion but also highlighted favorable features, including clear margins and no cancer in any of the 28 assessed lymph nodes. They received adjuvant radiation treatment. Recurrence was diagnosed 625 days after treatment completion. The last plasma sample for MRD detection was taken 554 days after treatment completion If their ctDNA levels had been monitored more regularly during the surveillance period, we posit that the recurrence may have been detected before reaching a level detectable by physical exam or cross-sectional imaging.

Patient SK_22 had a recurrent pT2N0M0 laryngeal tumor that was initially treated with radiation therapy three years prior to local recurrence. The patient underwent salvage total laryngectomy with bilateral cervical lymphadenectomy. Surgical pathology demonstrated lymphovascular invasion. The patient was not eligible for radiation therapy given their prior exposure. They ultimately developed a distant metastasis to the lung 191 days following treatment completion. Although SK_22’s postoperative tumor fractions were undetectable, their MAESTRO test may have been underpowered. SK_22’s last postoperative sample had the highest (i.e., worst) limit of detection and one of the lowest number of mutations assayed among all patents, which may have contributed to the lack of MRD detection (Figure 3B).

### Case studies: leveraging MRD detection to navigate therapeutic and diagnostic challenges

#### Escalation of adjuvant therapy in aggressive HNSCC

SK_47 was a patient with a recurrent pT2N1 oral cavity SCC who underwent salvage oral cavity composite resection and left cervical lymphadenectomy with reconstruction. Surgical pathology demonstrated extensive perineural invasion but no additional high-risk pathology. A multidisciplinary tumor board recommended adjuvant radiation to the immediate postoperative area to minimize overlap with prior irradiated fields and without chemotherapy consistent with the lack of extranodal extension or positive margins. MAESTRO testing was positive in the immediate postoperative window (postoperative day [POD] 1), prior to radiation initiation (POD 47) and following adjuvant radiotherapy on POD 131. Clinical recurrence was eventually identified on surveillance imaging on POD 173, a lead time of 5.8 months from molecular detection of recurrence to clinical detection of recurrence (Figure 4A). The patient received palliative immunotherapy for their recurrence but ultimately succumbed to their disease 210 days after surgery. This case highlights the opportunity for escalation of adjuvant therapy through either the addition of chemotherapy to adjuvant radiation had MAESTRO testing been utilized for clinical decision making, as MRD testing was persistently positive following surgery, or the early use of immunotherapy, as MAESTRO testing remained positive following radiation.

**Figure 4:**
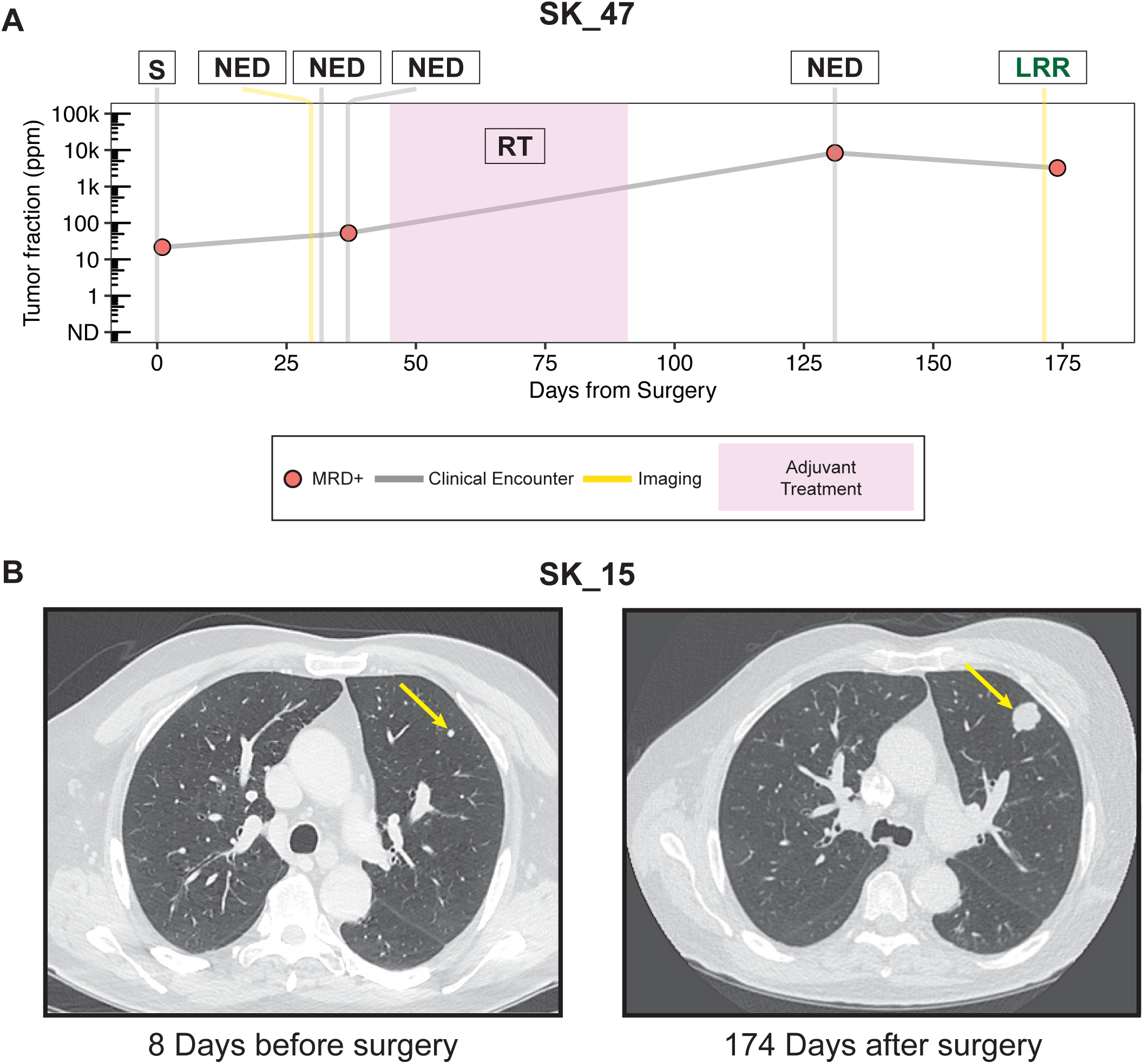
Clinical vignettes demonstrating how MRD detection could help navigate therapeutic and diagnostic challenges. (A) Longitudinal monitoring of tumor fraction levels in patient SK_47, with circles representing plasma sampling timepoints. Gray vertical lines mark clinical encounters where the patient was examined by an oncology provider, while yellow lines indicate instances of cross-sectional imaging. S = surgery, NED = no evidence of disease, RT = radiation therapy, LRR = locoregional recurrence. (B) An equivocal lung nodule identified 8 days before surgery deemed radiologically benign (yellow arrow; left panel), showed enlargement on surveillance imaging 174 days after surgery (right panel) and was confirmed to be a metastatic lesion 203 days after surgery.

#### Synchronous equivocal lung nodules

Patient SK_15 had a 3 mm lung nodule found on initial staging scans that was noted to likely be benign but warranted follow up as determined by the radiologist and multidisciplinary tumor board review (Figure 4B). They were recommended for curative-intent resection of pT4aN3bM0 oral cavity SCC. Following surgery, MAESTRO testing remained positive throughout the post-operative window (Figure 1A). The patient underwent pathology-guided adjuvant chemoradiotherapy due to evidence of extranodal extension on surgical pathology as determined by a multidisciplinary tumor board. On 6-month surveillance imaging, the patient was found to have an interval increase in size of the pre-existing lung nodule (Figure 4B). Biopsy of the lung nodule demonstrated SCC. They were started on chemoimmunotherapy and received stereotactic body radiotherapy to the lung nodule. Despite these treatments, the patient ultimately succumbed to the disease and died 723 days after treatment completion. In this case, we hypothesize that the persistent MRD after surgery and chemoradiotherapy was due to the metastatic lung nodule that was present at the time of initial surgery yet not recognized by standard imaging-based approaches.

## Discussion

In this study, we demonstrate the application of MAESTRO for MRD detection in the immediate postoperative setting to predict recurrence and death in a cohort of predominantly advanced-stage, HPV-independent HNSCC patients. To our knowledge, these results are the first to show that MRD detection as early as the first 1-3 postoperative days can be a highly sensitive and specific predictor of recurrence and death in HPV-independent HNSCC patients.

Several studies have explored tumor-informed detection of ctDNA in cohorts of HPV-independent HNSCC patients. The LIONESS study involved a cohort of 17 patients who underwent postoperative ctDNA testing utilizing the RaDaR^TM^ assay (30). In this study, ctDNA was detected before clinical progression in all 5 patients who experienced progression, although the study did not evaluate the effectiveness of immediate postoperative sampling in predicting survival outcomes. In a similar but larger study, Hanna *et al.* applied the Signatera assay in 14 HPV-independent surgically treated HNSCC patients with the first post-surgical test conducted at a median of 24 days after surgery (22). The Signatera assay is known to have a limit of detection of 100 ppm (22). Our findings indicate that the majority of early postoperative MRD-positive samples had tumor fraction levels below this threshold, highlighting the need for an ultrasensitive assay like MAESTRO to track MRD accurately at timepoints that are clinically relevant for clinical decision-making. Adjuvant clinical decision making in HNSCC occurs 1-2 weeks after surgery, so that adjuvant treatment can begin within a 6-week window, which has been shown to impact survival (31). Thus, MRD timepoints used in many other cancer types, which occur weeks after surgery, are not as clinically relevant in HNSCC. In this study, we show that ultra-early MRD sampling, conducted while patients remain in the hospital after surgery, is feasible and efficacious, so that results are returned to providers within a clinically relevant timeframe for adjuvant decision making. While immediate post-operative MRD sampling has been studied in other cancer types, including HPV-associated HNSCC and nasopharyngeal cancer, no such studies have been conducted in HPV-independent HNSCC (32,33)

Others have highlighted the advantages of tumor-agnostic MRD testing methods. A recent study demonstrated a tumor-agnostic assay using a 26-gene panel of commonly mutated genes in HNSCC along with 2 HPV16 genes that could detect pre-treatment ctDNA in 77% of patients and predict disease progression (34). Yet, tumor agnostic assays often cannot detect below 100 ppm ctDNA (35,36). In another recent study, a tissue agnostic genome-wide methylome enrichment MRD assay called cfMeDIP-seq was used for MRD detection in patients with HNSCC but did not assess its clinical performance in the immediate post-treatment setting (37).

In our clinical scenarios of selected patients, the two patients with false-negative tests underscore the importance of (1) implementing more frequent monitoring of ctDNA levels during the surveillance period, as demonstrated by patient SK_07, and (2) maintaining a critical awareness of the technical sensitivity of each patient’s bespoke assay to ensure its reliability for clinical decision-making, as exemplified by SK_22. These patients with false-negative tests may have benefitted from using larger plasma volumes in order to increase the sensitivity and robustness of ctDNA testing (26). In the case of patient SK_48, who initially had a false-positive result based on the first postoperative sample, their eventual clearance of tumor fraction levels suggests the presence of impaired cfDNA clearance kinetics. One of the major mechanisms of natural cfDNA clearance is uptake by liver-resident macrophages, Kupffer cells (38–40). Interestingly, this patient had a documented history of nonalcoholic fatty liver disease (NAFLD), a condition known to alter the liver microenvironment and impair the phagocytic function of Kupffer cells (41,42). This raises the possibility that SK_48’s NAFLD contributed to delayed ctDNA clearance, resulting in a slower clearance of tumor fraction levels until the last postoperative sample, 7 days after surgery.

As EFS and OS were significantly inferior for patients with MRD positivity, our data suggests that ultra-early MRD detection after surgery can identify high-risk HNSCC patients with markedly worse survival, highlighted by two patients who may have benefitted from MRD-informed clinical decision-making. For example, patient SK_47’s persistently positive MRD status following surgery could have been used to intensify adjuvant therapy with the addition of chemotherapy despite not meeting conventional pathologic criteria. In the case study of patient SK_15, we demonstrated how MRD detection can address a significant diagnostic and therapeutic challenge in managing HNSCC populations: discerning the nature of lung nodules of undetermined significance, a frequent occurrence (43). Many HNSCC patients with a history of former or active smoking frequently present with lung nodules that do not initially meet radiologic criteria for pathologic disease on staging scans (44,45). Such nodules are often categorized as probably benign in the absence of obvious radiologic findings but warranting clinical follow up. Persistent ctDNA detection following surgery could have led to a higher degree of concern for this suspicious nodule, which, in turn, could have been used to intervene earlier with the potential to increase the chance of cure.

A limitation of this study was the non-standardized sampling times in the surveillance period. Despite this, the relatively long median follow-up time of our study of nearly 2 years relative to other prospective MRD studies captures a high-risk time period in HNSCC surveillance as most recurrences (around 80%) occur within this timeframe (46). The inclusion of only one p16-positive HNSCC patient limits the generalizability of the findings to this disease subtype, but our whole-genome, tumor-informed approach through MAESTRO-Pool should be applicable to either etiology of HNSCC. Given that our providers were blinded to the MRD results, it remains to be determined how MRD-informed clinical decision making could impact patient outcomes. Moving forward, thoughtfully designed randomized trials are warranted that include ultrasensitive MRD detection testing of patients in the immediate postoperative phase for risk-adapted clinical decision making in HNSCC.

## Data Availability

All data produced in the present study are available upon reasonable request to the authors

## Acknowledgements

The authors acknowledge the generous support of the Gerstner Family Foundation and funding support from the NIH National Cancer Institute (2R01CA221874-04). S.T. acknowledges funding support from NIH K08EB036081.

## Supplemental Material

Supplemental material is available online

