## Supplemental Figures for "Immediate postoperative minimal residual disease detection with MAESTRO predicts recurrence and survival in head and neck cancer patients treated with surgery"

### Supplemental Figure 1

| Variable | | N | Hazard ratio | | p |
|---|---|---|---|---|---|
| **Age at Surgery** | | 24 | | 0.97 (0.92, 1.02) | 0.206 |
| **Smoking History** | Never | 6 | | Reference | |
| | Ever | 18 | | 3.17 (0.71, 14.12) | 0.130 |
| **Overall Pathologic Stage** | | 24 | | 2.14 (0.86, 5.36) | 0.103 |
| **Primary Site** | Oral Cavity | 19 | | Reference | |
| | Larynx/Hypopharynx | 3 | | 1.45 (0.31, 6.74) | 0.634 |
| | Oropharynx | 2 | | 3.92 (0.79, 19.46) | 0.095 |
| **First Postop Sample** | MRD- | 10 | | Reference | |
| | MRD+ | 14 | | 20.42 (2.61, 159.56) | 0.004 |
| **Last Postop Sample** | MRD- | 11 | | Reference | |
| | MRD+ | 13 | | 27.36 (3.49, 214.47) | 0.002 |
| **Perineural Invasion** | | 24 | | 1.96 (0.44, 8.80) | 0.378 |
| **Lymphovascular Invasion** | | 24 | | 3.13 (1.02, 9.58) | 0.046 |
| **Extranodal Extension and/or Positive Margin** | | 24 | | 3.39 (1.10, 10.42) | 0.033 |

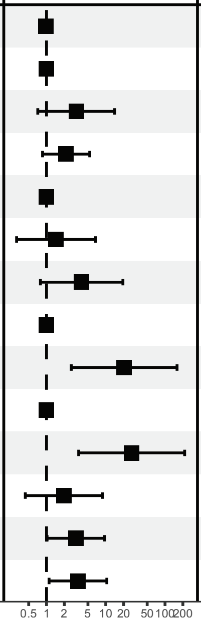

### Supplemental Figure 2

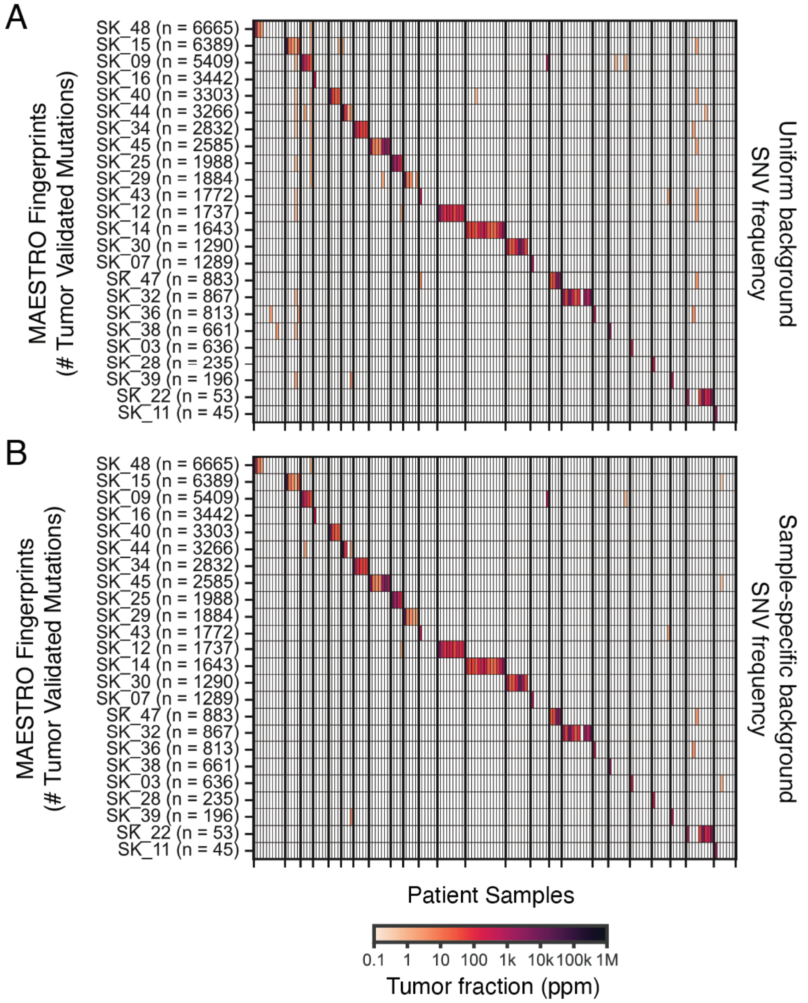


C

| | MRD+ Calls | |
|---|---|---|
| | Uniform SNV Frequency | Sample-Specific SNV Frequencies |
| Matched Tests | 97 / 155 | 97* / 155 |
| Unmatched Tests | 42 / 3,565 | 12 / 3,565 |

*SK_29 day 374 changes to MRD+, SK_44 day 3 changes to MRD-

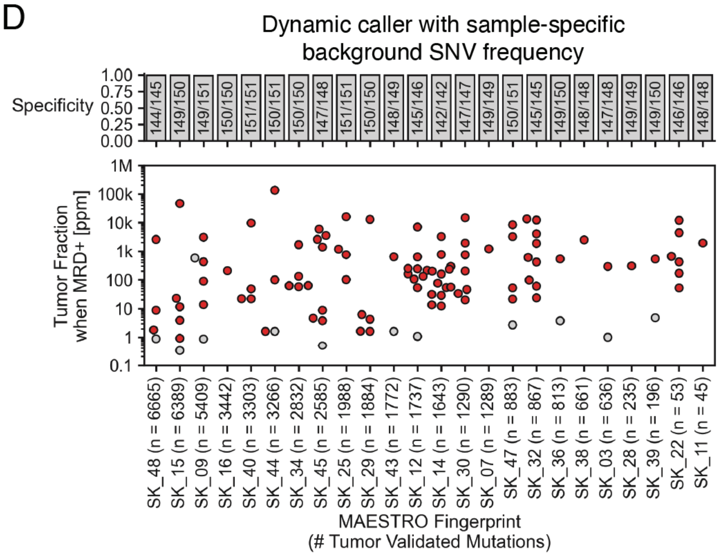

**Supplemental Figure 3**

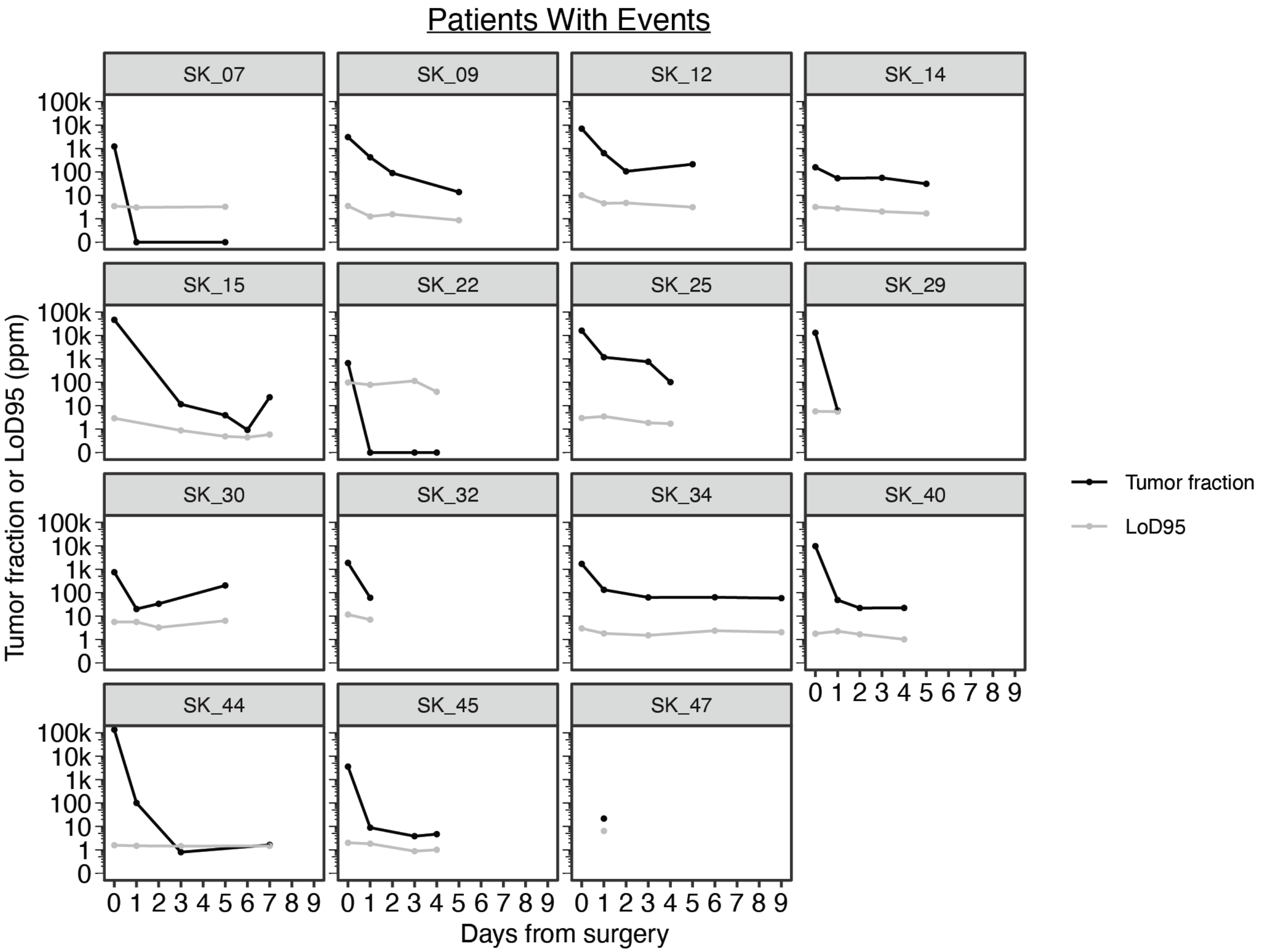


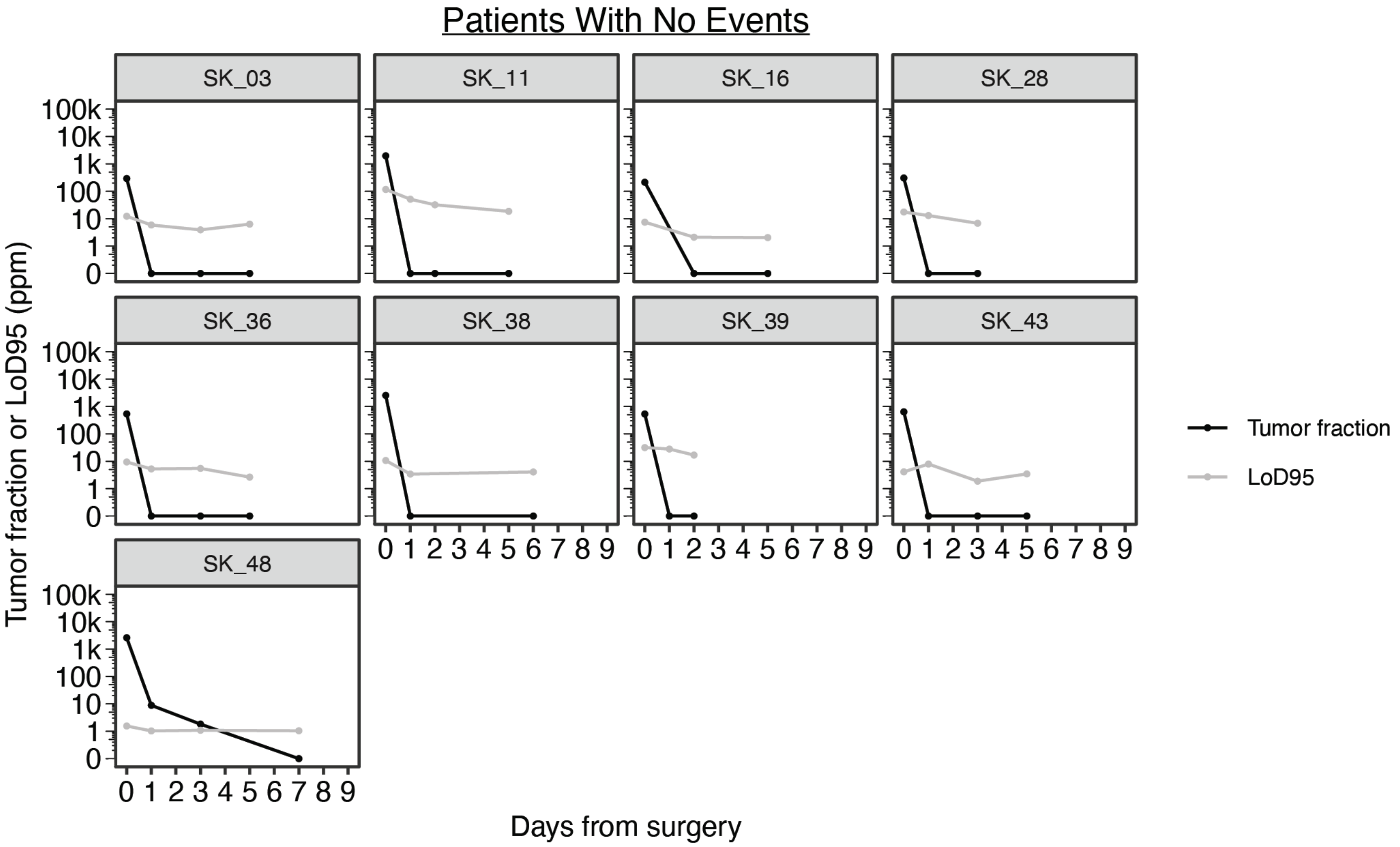
